# Plasma neurofilament light levels show elevation prior to diagnosis of sporadic motor neuron disease in the UK Biobank cohort

**DOI:** 10.1101/2023.02.27.23286529

**Authors:** Erin N. Smith, Jonghun Lee, Daria Prilutsky, Stephen Zicha, Zemin Wang, Steve Han, Neta Zach

## Abstract

**Objective:** Motor neuron disease (MND) is a debilitating neurodegenerative disease with profound unmet need. In pre-symptomatic mutation carriers, elevations in neurofilament light (NfL) precede symptom onset, however, the presence and timing of elevation is much more difficult to study in sporadic cases.

**Methods:** Using the UK Biobank cohort, we tested whether plasma NfL predicted risk of diagnosis of sporadic MND using survival analysis.

**Results:** We identified 241 MND patients with pre-diagnosis NfL data, of which 203 (84%) lacked predicted loss of function or deleterious missense variants in established ALS genes. A total of 42,752 controls without MND were obtained from a random sample of UK Biobank participants. At two years pre-diagnosis, we found that NfL levels in patients exceeded the 95^th^ percentile of controls and that patients could be discriminated from controls at high accuracy (AUC = 0.95 (95% CI 0.89-1.01)). In participants with hospital record follow-up after study enrollment, a 2-fold increase in NfL levels was associated with a 3.4 fold risk of receiving an MND diagnosis per year (95% CI 2.9-3.9, P = 4 × 10^−64^)).

**Discussion:** Our findings show that NfL can identify sporadic MND as early as 2 years prior to diagnosis.

## Introduction

Amyotrophic lateral sclerosis, also known as motor neuron disease (MND), is a neurodegenerative disease of motor neurons in the brain and spinal cord, leading to progressive weakness, paralysis and death, and the need for an effective therapy is profound. Among the challenges limiting the successful development of treatments is the limited understanding and ability to reliably track disease mechanisms early in disease onset or pre-symptomatically. Looking at carriers of genetic mutations leading to neurodegeneration suggests disease pathology starts decades before symptoms begin to appear^1, 2^, and understanding pre-symptomatic progression is critical to enable potential disease prevention interventions^1^.

Neurofilament light chain (NfL) is one of three types of intermediate filaments that are exclusively expressed in neurons^2, 3^. As axonal and neuronal death occurs, NfL has been consistently found to be elevated in CSF, plasma and serum of patients with neurodegenerative diseases such as MND, Huntington’s disease, multiple sclerosis and others^2, 4, 5^. In MND, absolute levels of NfL predict future survival and strongly and reliably relate to clinical progression early in the disease^6^. NfL has been shown to be elevated in pre-symptomatic mutation carriers as they approach symptom onset^7-9^. Early data suggests similar trend in broad ALS patient population^10^. However, the extent and timeline to which pre-symptomatic elevation occurs in the broader sporadic disease remains to be elucidated.

Understanding the natural history of NfL change in pre-symptomatic MND patients and its relationship with symptomatic and diagnostic conversion is highly needed to better understand disease mechanisms, aid in the earlier identification of patients-at-risk, and decrease the diagnostic delay to maximize the potential of early intervention of future disease modifying therapies. However, given that MND is a rare mostly sporadic disease, large population-based studies are required, including both familial and sporadic MND. Recently, such a cohort became a reality through the creation of the UK Biobank^11^, including 500,000 participants with clinical and genetic data, and blood samples. Recently, the UK Biobank Pharma Proteomics Project (UKB-PPP) has generated proteomics data from the plasma samples of 54,306 UKB participants^12^, enabling the analysis of NfL levels prior to a diagnosis of MND.

Here, we examine whether NfL is predictive of developing sporadic MND by performing a survival analysis of NfL levels obtained prior to MND diagnosis in 203 individuals who did not carry a predicted deleterious variant in a risk gene for amyotrophic lateral sclerosis compared to 42,752 controls. To our knowledge this is the first comprehensive natural history study of pre-symptomatic changes in sporadic MND and suggests a novel path to investigate early disease mechanism and intervention studies.

## Methods

Full Methodology is available in the Supplementary Methods. Briefly,

This research has been conducted using the UK Biobank^11^ Resource (Application Numbers 26041 and 65851). Subjects were selected from the UK Biobank Pharma Proteomics Project (UKB-PPP)^12^. Cases were selected from those that received a hospital record diagnosis of G12.2 Motor Neuron Disease. Additionally, cases that did not carry a predicted deleterious variant in genes that have been implicated in familial ALS^13^ were considered sporadic (suppl. Methods and Table S2). The time of first diagnosis was obtained from hospital record data (see also Table S1). NfL levels were measured using the NEFL assay (OID20871) from the Olink Explore 1536 panel as part of the UKB-PPP. Survival analysis was performed using Cox proportional hazards regression (with and without covariates: sex, age at recruitment, fasting time, years between plasma sample collection and protein analysis, and ethnicity).

## Results

Participant characteristics are summarized in Table 1. Briefly, 241 individuals with available plasma proteomics samples were identified based on hospital records of MND (G12.2 diagnosis) occurring after study enrollment. Patients with hospital diagnosis, a general practitioner diagnosis, or a self-report of MND at study entry, were excluded. As family history of motor neuron disease was not available, analysis of ALS associated genes^13^ (excluding the C9orf72 hexanucleotide repeat which cannot be ascertained with exome sequencing, Table S1) identified 38 individuals, resulting in 203 without known exonic mutations that were considered sporadic. Following that, a control group of 42,752 participants without a G12.2 diagnosis was selected from the randomly selected cohort of UKB-PPP participants.

**Table 1.** Participant characteristics

|  | Cases | Controls |
| --- | --- | --- |
| **N** | 203 | 42752 |
| **Years of age at enrollment (mean (sd))** | 60.1 (7.0) | 56.7 (8.1) |
| **Female Sex (N (%))** | 94 (46.3%) | 23174 (54.2%) |
| **NfL levels (NPX) (mean (sd))** | 0.6 (0.9) | 0 (0.5) |
| **Years of age at onset (mean (sd))** | 65.6 (7.4) | -- |
| **Years prior to diagnosis (mean (sd))** | 5.6 (2.8) | -- |
| **Number of cases that died from G12.2** | 136 | -- |
| **Years from diagnosis to death** | 1.6 (1.2) | -- |
| **Self-reported race** |  |  |
| Asian or Asian British | 1 (0.5%) | 837 (2.0%) |
| Black or Black British | 2 (1.0%) | 722 (1.7%) |
| Chinese | - | 132 (0.3%) |
| Mixed | 3 (1.5%) | 282 (0.7%) |
| Other ethnic group | - | 412 (1.0%) |
| White | 195 (96.1%) | 40157 (93.9%) |
| Not reported | 2 (1.0%) | 210 (0.5%) |

We examined the relationship between NfL levels and MND using plasma levels from the UKB-PPP (Figure 1). NfL levels were 1.54-fold higher in sporadic patients subsequently diagnosed with motor neuron disease (Figure 1A; P = 2.3 × 10^−58^). Results adjusted for age, sex, fasting time, length of plasma storage, and ethnic group were similar (1.41 fold-higher, P = 7 × 10^−50^). When examined relative to the time of diagnosis, NfL levels were highest when measured within 2-3 years prior to diagnosis, and exceeded the 95% percentile of controls more than 2 years prior to diagnosis (Figure 1B). A survival analysis of 201 cases and 31,253 controls showed a 3.4 increased risk of a MND diagnosis per year for each 2-fold increase (1 NPX unit) in NfL (HR = 3.4 (95% CI 2.9-3.9, P = 4 × 10^−64^)) with similar results when adjusted for age, sex, fasting time, length of plasma storage, and ethnic group (HR = 3.4, P = 2 × 10^−53^). The predictive accuracy of NfL was high, and NfL was singlehandedly predictive of diagnosis 2 years prior, with an area under the curve (AUC) of 0.95 (95% CI 0.89-1.01) (Figure 2).

**Figure 1:**
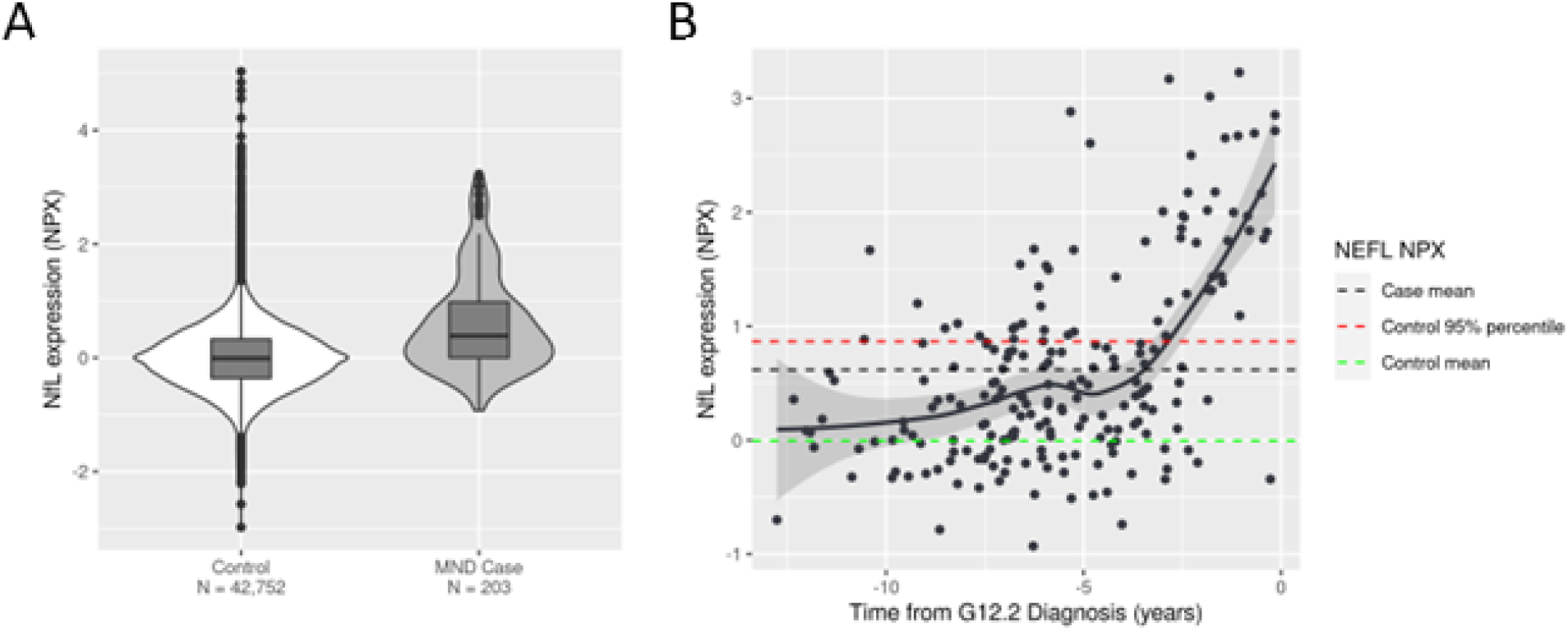
NfL levels are elevated in sporadic MND cases and increase relative to time of diagnosis. NfL expression level is reported as Normalized Protein eXpression (NPX) as measured by the NEFL Olink Explore 1536 assay, with each unit corresponding to a 2-fold difference. A) Violin and boxplot showing the distribution of NfL levels in MND cases vs. controls. B) Scatterplot showing NfL expression vs. time from motor neuron disease diagnosis in 203 cases. The black solid line indicates a smoothed line calculated using the loess method. Dotted lines indicate average, or 95% percentile levels of NfL expression for the indicated groups.

**Figure 2:**
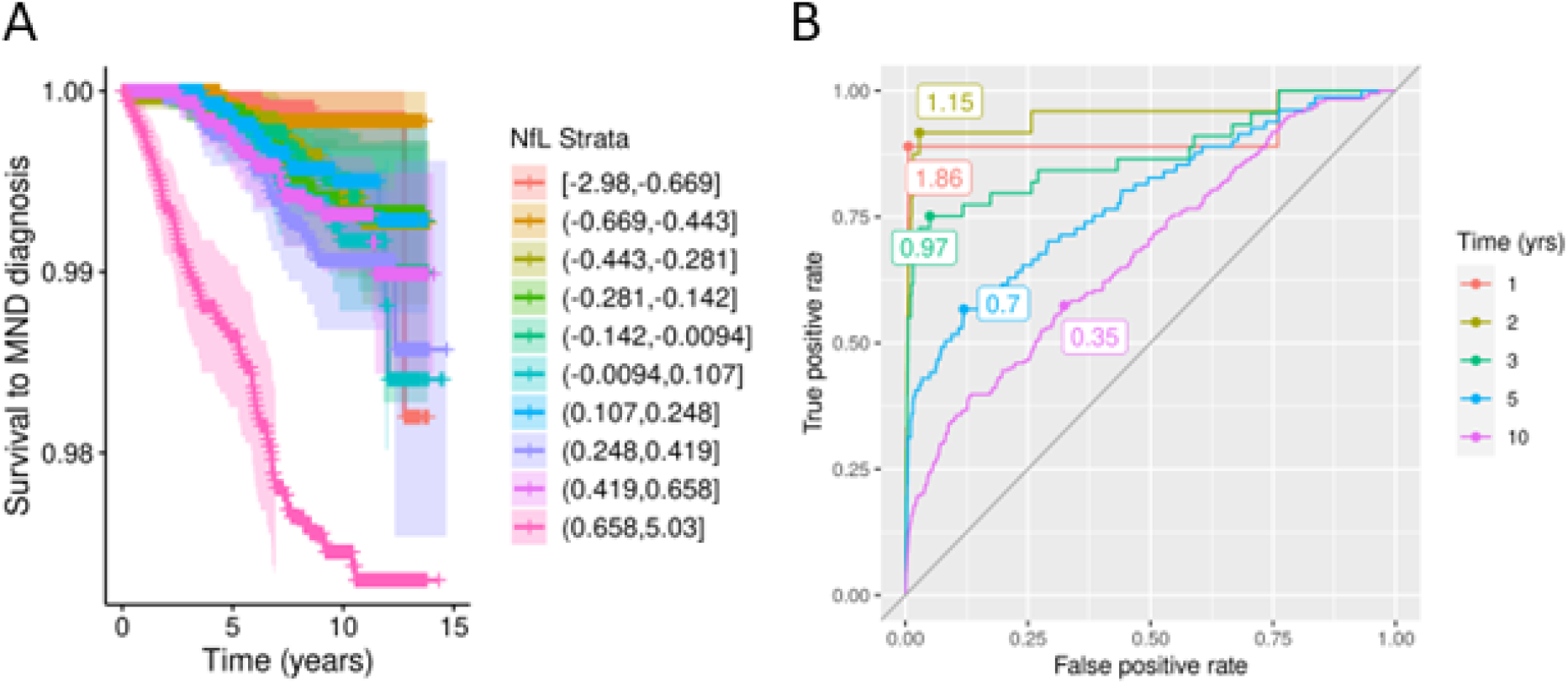
NfL significantly predicts time to MND diagnosis in 201 cases and 31,253 controls. A) Kaplan-Meier plot showing the probability of participants developing an MND diagnosis by time. Subjects are grouped into strata by decile of NfL expression. Horizonal tick marks indicate censored subjects. B) Time-dependent receiver-operator curves showing the relationship between true and false positive rates at specific time points. The numbers indicate the NfL NPX values that optimally balance true positive and false positive rates.

The pattern of NfL changes was similar between patients with and without identified ALS mutations (Figure S1, Table S1). However, NfL changes in patients pre-symptomatic to a diagnosis with either Parkinson’s disease, Alzheimer’s disease, or circumscribed brain atrophy (which includes frontotemporal dementia), did not show the same strong elevation prior to diagnosis (Figure S3). While NfL was elevated in these diseases before diagnosis, it showed weaker predictive accuracy relative to MND (Table S3), potentially reflecting the fast progressive nature of MND. In addition, we explored whether hand-grip strength, a symptom occurring early in limb-onset ALS, was predictive of time to diagnosis and did not observe significant association (Figure S3;), suggesting that NfL is a stronger predictor of diagnosis.

## Discussion

Overall, we observed in a large cohort that NfL was significantly elevated compared to controls prior to MND diagnosis (on average 1.54-fold higher) with a 2-fold increase in NfL being associated with a 3.4-fold increase in risk of MND per year. NfL was a reliable predictor of diagnosis with a peak time-dependent AUC of 95% at two years before diagnosis. To date, this study represents the largest exploration of NfL change in pre-symptomatic sporadic MND.

These results are similar in nature to those reported by the pre-fALS study of mutation carriers^8, 9^ and the general MND population^10^, and expand them to sporadic patients, as well as a larger cohort of familial patients. Considering a diagnostic delay of 11 months^14^, these results suggest that neurodegeneration occurs prior to symptomatic manifestation and that NfL could aid in early diagnosis of MND, and early interventions by therapeutic agents.

This analysis of pre-symptomatic sporadic patients would not have been made possible without the thorough design of the UK Biobank study. However, there are a few limitations to note-first, the UK Biobank data doesn’t contain detailed neurological examination at patient recorded symptom onset and therefore precise time prior to symptom onset cannot be assessed. Second, data analyzed here is cross sectional, with each patient providing a single plasma sample, and individual longitudinal changes and covariates are not well explored. Third, the limited availability of whole genome sequence, precluded the identification of C9orf72 hexanucleotide repeat mutation carriers. Finally, as the UK Biobank is predominantly of White ancestry, additional studies of diverse populations would be needed to generalize the results.

## Data Availability

All data used in the present study will be available through the UK Biobank.

## Acknowledgements

The authors would like to thank the participants of the UK Biobank study for enabling this work.

## Disclosure of relationships and activities

All authors are employees of and own stock in Takeda Development Center Americas, Inc.

---

STROBE Statement—checklist of items that should be included in reports of observational studies

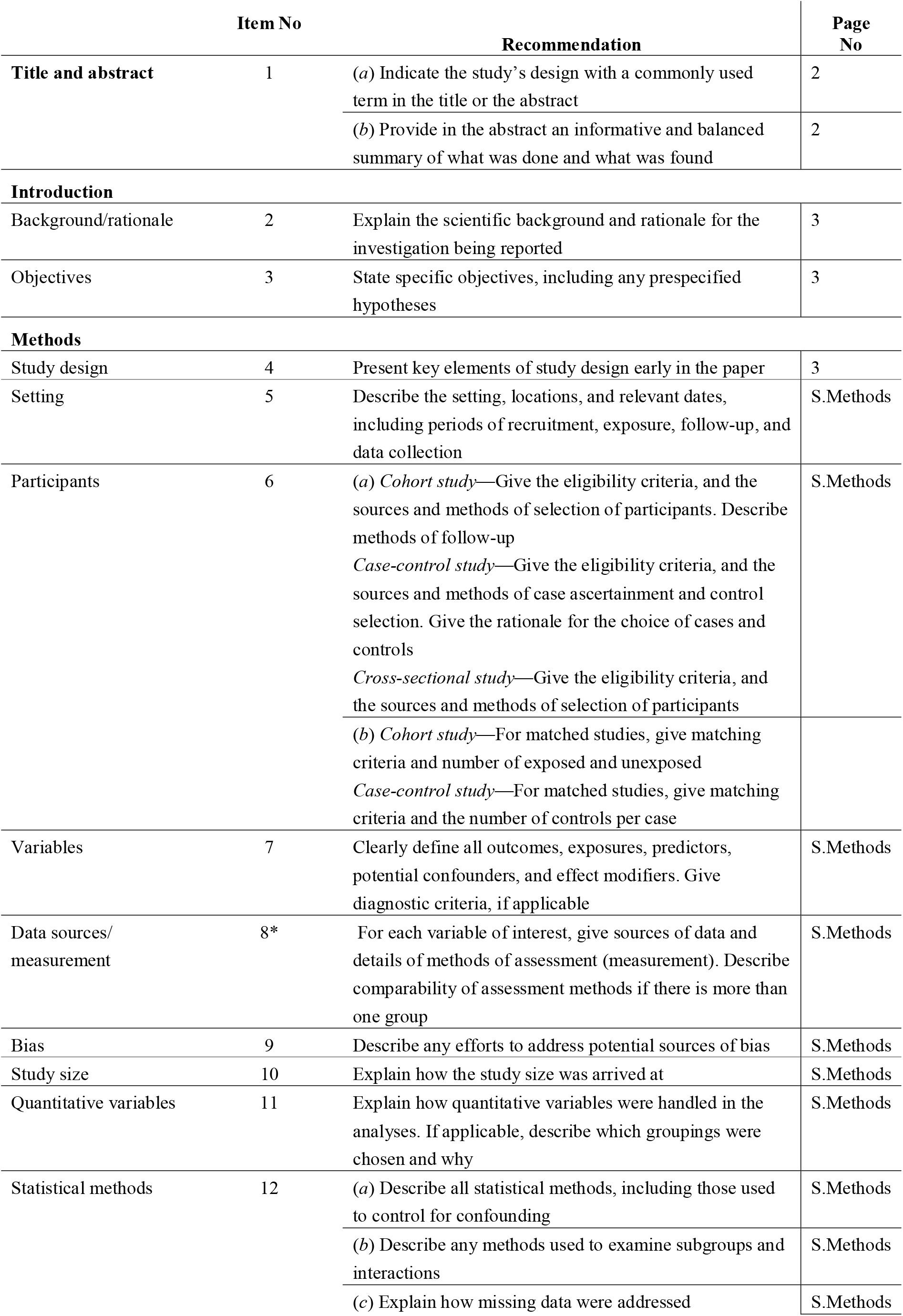
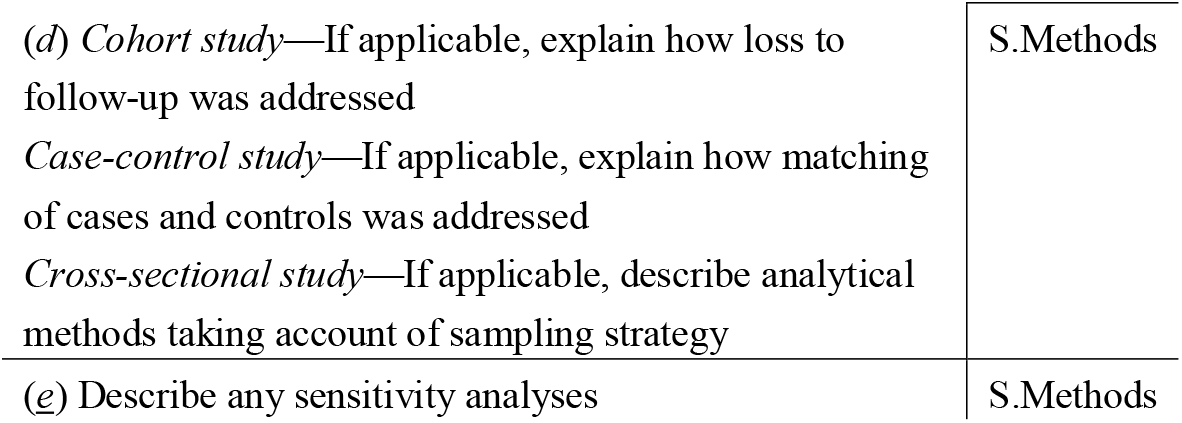

|  | Item No | Recommendation | Page No |
| --- | --- | --- | --- |
| Title and abstract | 1 | (a) Indicate the study’s design with a commonly used term in the title or the abstract | 2 |
|  |  | (b) Provide in the abstract an informative and balanced summary of what was done and what was found | 2 |
| Introduction |  |  |  |
| Background/rationale | 2 | Explain the scientific background and rationale for the investigation being reported | 3 |
| Objectives | 3 | State specific objectives, including any prespecified hypotheses | 3 |
| Methods |  |  |  |
| Study design | 4 | Present key elements of study design early in the paper | 3 |
| Setting | 5 | Describe the setting, locations, and relevant dates, including periods of recruitment, exposure, follow-up, and data collection | S.Methods |
| Participants | 6 | (a) Cohort study—Give the eligibility criteria, and the sources and methods of selection of participants. Describe methods of follow-up<br>Case-control study—Give the eligibility criteria, and the sources and methods of case ascertainment and control selection. Give the rationale for the choice of cases and controls<br>Cross-sectional study—Give the eligibility criteria, and the sources and methods of selection of participants | S.Methods |
|  |  | (b) Cohort study—For matched studies, give matching criteria and number of exposed and unexposed<br>Case-control study—For matched studies, give matching criteria and the number of controls per case |  |
| Variables | 7 | Clearly define all outcomes, exposures, predictors, potential confounders, and effect modifiers. Give diagnostic criteria, if applicable | S.Methods |
| Data sources/<br>measurement | 8* | For each variable of interest, give sources of data and details of methods of assessment (measurement). Describe comparability of assessment methods if there is more than one group | S.Methods |
| Bias | 9 | Describe any efforts to address potential sources of bias | S.Methods |
| Study size | 10 | Explain how the study size was arrived at | S.Methods |
| Quantitative variables | 11 | Explain how quantitative variables were handled in the analyses. If applicable, describe which groupings were chosen and why | S.Methods |
| Statistical methods | 12 | (a) Describe all statistical methods, including those used to control for confounding | S.Methods |
|  |  | (b) Describe any methods used to examine subgroups and interactions | S.Methods |
|  |  | (c) Explain how missing data were addressed | S.Methods |

|  |  |
| --- | --- |
| (d) *Cohort study*—If applicable, explain how loss to follow-up was addressed<br>*Case-control study*—If applicable, explain how matching of cases and controls was addressed<br>*Cross-sectional study*—If applicable, describe analytical methods taking account of sampling strategy | S.Methods |
| (e) Describe any sensitivity analyses | S.Methods |

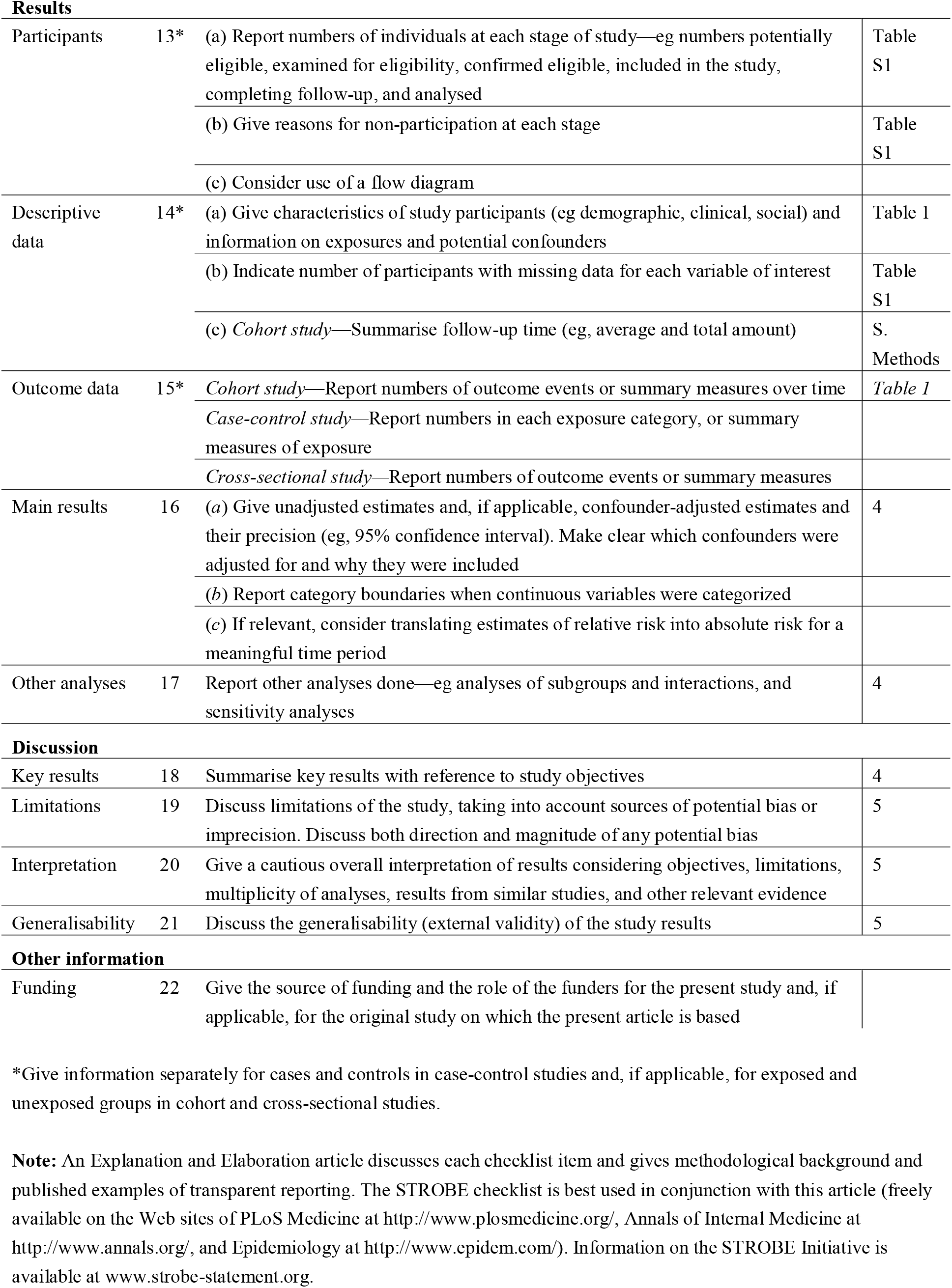

|  |  |  |  |
| --- | --- | --- | --- |
| Participants | 13* | (a) Report numbers of individuals at each stage of study—eg numbers potentially eligible, examined for eligibility, confirmed eligible, included in the study, completing follow-up, and analysed | Table S1 |
|  |  | (b) Give reasons for non-participation at each stage | Table S1 |
|  |  | (c) Consider use of a flow diagram |  |
| Descriptive data | 14* | (a) Give characteristics of study participants (eg demographic, clinical, social) and information on exposures and potential confounders | Table 1 |
|  |  | (b) Indicate number of participants with missing data for each variable of interest | Table S1 |
|  |  | (c) *Cohort study*—Summarise follow-up time (eg, average and total amount) | S. Methods |
| Outcome data | 15* | *Cohort study*—Report numbers of outcome events or summary measures over time | Table 1 |
|  |  | *Case-control study*—Report numbers in each exposure category, or summary measures of exposure |  |
|  |  | *Cross-sectional study*—Report numbers of outcome events or summary measures |  |
| Main results | 16 | (a) Give unadjusted estimates and, if applicable, confounder-adjusted estimates and their precision (eg, 95% confidence interval). Make clear which confounders were adjusted for and why they were included | 4 |
|  |  | (b) Report category boundaries when continuous variables were categorized |  |
|  |  | (c) If relevant, consider translating estimates of relative risk into absolute risk for a meaningful time period |  |
| Other analyses | 17 | Report other analyses done—eg analyses of subgroups and interactions, and sensitivity analyses | 4 |
| Key results | 18 | Summarise key results with reference to study objectives | 4 |
| Limitations | 19 | Discuss limitations of the study, taking into account sources of potential bias or imprecision. Discuss both direction and magnitude of any potential bias | 5 |
| Interpretation | 20 | Give a cautious overall interpretation of results considering objectives, limitations, multiplicity of analyses, results from similar studies, and other relevant evidence | 5 |
| Generalisability | 21 | Discuss the generalisability (external validity) of the study results | 5 |

Other information
|  |  |  |
| --- | --- | --- |
| Funding | 22 | Give the source of funding and the role of the funders for the present study and, if applicable, for the original study on which the present article is based |

\*Give information separately for cases and controls in case-control studies and, if applicable, for exposed and unexposed groups in cohort and cross-sectional studies.
**Note:** An Explanation and Elaboration article discusses each checklist item and gives methodological background and published examples of transparent reporting. The STROBE checklist is best used in conjunction with this article (freely available on the Web sites of PLoS Medicine at http://www.plosmedicine.org/, Annals of Internal Medicine at http://www.annals.org/, and Epidemiology at http://www.epidem.com/). Information on the STROBE Initiative is available at www.strobe-statement.org.

## Notes

### Funding Statement

This study did not receive any funding

### Author Declarations

This research has been conducted using the UK Biobank Resource under Application Numbers 26041 and 65851. The UK Biobank was approved by the North West Multi-centre Research Ethics Committee (MREC) as a Research Tissue Bank (RTB) approval, which allows use by approved researchers without additional ethical clearance.

## References

1. Panza F, Seripa D, Lozupone M, et al. The potential of solanezumab and gantenerumab to prevent Alzheimer’s disease in people with inherited mutations that cause its early onset. Expert Opin Biol Ther 2018;18:25–35.

2. Bacioglu M, Maia LF, Preische O, et al. Neurofilament Light Chain in Blood and CSF as Marker of Disease Progression in Mouse Models and in Neurodegenerative Diseases. Neuron 2016;91:56–66.

3. Friede RL, Samorajski T. Axon caliber related to neurofilaments and microtubules in sciatic nerve fibers of rats and mice. Anat Rec 1970;167:379–387.

4. Gaetani L, Blennow K, Calabresi P, Di Filippo M, Parnetti L, Zetterberg H. Neurofilament light chain as a biomarker in neurological disorders. J Neurol Neurosurg Psychiatry 2019;90:870–881.

5. Olsson B, Portelius E, Cullen NC, et al. Association of Cerebrospinal Fluid Neurofilament Light Protein Levels With Cognition in Patients With Dementia, Motor Neuron Disease, and Movement Disorders. JAMA Neurol 2019;76:318–325.

6. Lu CH, Macdonald-Wallis C, Gray E, et al. Neurofilament light chain: A prognostic biomarker in amyotrophic lateral sclerosis. Neurology 2015;84:2247–2257.

7. Benatar M, Wuu J. Presymptomatic studies in ALS: rationale, challenges, and approach. Neurology 2012;79:1732–1739.

8. Benatar M, Wuu J, Andersen PM, Lombardi V, Malaspina A. Neurofilament light: A candidate biomarker of presymptomatic amyotrophic lateral sclerosis and phenoconversion. Ann Neurol 2018;84:130–139.

9. Benatar M, Wuu J, Lombardi V, et al. Neurofilaments in pre-symptomatic ALS and the impact of genotype. Amyotroph Lateral Scler Frontotemporal Degener 2019;20:538–548.

10. Bjornevik K, O’Reilly EJ, Molsberry S, et al. Prediagnostic Neurofilament Light Chain Levels in Amyotrophic Lateral Sclerosis. Neurology 2021;97:e1466–1474.

11. Ollier W, Sprosen T, Peakman T. UK Biobank: from concept to reality. Pharmacogenomics 2005;6:639–646.

12. Sun BB, Chiou J, Traylor M, et al. Genetic regulation of the human plasma proteome in 54,306 UK Biobank participants. bioRxiv 2022.

13. Mejzini R, Flynn LL, Pitout IL, Fletcher S, Wilton SD, Akkari PA. ALS Genetics, Mechanisms, and Therapeutics: Where Are We Now? Front Neurosci 2019;13:1310.

14. Paganoni S, Macklin EA, Lee A, et al. Diagnostic timelines and delays in diagnosing amyotrophic lateral sclerosis (ALS). Amyotroph Lateral Scler Frontotemporal Degener 2014;15:453–456.

