## Supplementary materials for "Plasma neurofilament light levels show elevation prior to diagnosis of sporadic motor neuron disease in the UK Biobank cohort"

**Supplementary Methods**

*Subject selection*

This research has been conducted using the UK Biobank Resource under Application Numbers 26041 and 65851. The UK Biobank was approved by the North West Multi-centre Research Ethics Committee (MREC) as a Research Tissue Bank (RTB) approval, which allows use by approved researchers without additional ethical clearance. Participants provided informed consent^1^. Subjects were selected from those in the UK Biobank study^1^ where proteomics data were generated as part of the UK Biobank Pharma Proteomics Project (UKB-PPP)^2^. Participants enrolled in the study from 2006-2010 and linked health record data was obtained through 2021. Cases and controls had average follow-up times of 5.6 years and 8.4 years, respectively. Cases were selected from those that received a hospital record diagnosis of G12.2 Motor Neuron Disease. Participants were excluded if they received a G12.2 diagnosis prior to study entry, self-reported having a motor neuron disease at study entry (Field 20002-0), or a motor neuron disease diagnosis (READ2/3 codes F15.., F152., F1521, F1522, F1523, F1524, and F152z; or READ3 codes X00A1 and XE17v) from their general practitioner prior to study entry. Additionally, cases that did not carry a predicted deleterious variant in genes that have been implicated in familial ALS^3^ were considered sporadic. The time of first diagnosis was obtained from hospital record data. A total of 241 MND patients with pre-diagnosis NfL data, of which 203 (84%) lacked predicted loss of function or deleterious missense variants in established ALS genes, and 42,752 controls were identified. Detailed counts of subjects filtered by different factors are presented in Table S1. For the survival analysis, follow-up time was considered as the time from study enrollment to the last date in the hospital record data. Of the 203 cases and 42,752 controls, 201 cases and 31,253 controls that had at least one hospital record diagnosis after study entry and complete covariate data were included in the survival analysis.

*Proteomics*

NfL levels were measured using the NEFL assay (OID20871) from the Olink Explore 1536 panel as part of the UKB-PPP. Data from baseline samples (Instance 0) from subjects that were part of the Random group, or the Consortium group with a diagnosis of G12.2, was included in the analysis. Measures with a non-missing measurement, and a QC_Warning and Assay_Warning of PASS were included, and additional normalization was not performed. Overall, 99.4% of samples showed levels greater than the limit of detection (LOD), and the data was not filtered based on this threshold.

*Exome data*

Genetic variants in 44 genes implicated in ALS^3^ were examined for potential deleterious variants using the 450K release of the UK Biobank exome sequencing (N = 454,775). Variant effect annotation was performed using Variant Effect Predictor (VEP) and the Loss-Of-Function Transcript Effect Estimator (LOFTEE) Plugin. Missense variants were considered deleterious if they had a predicted pathogenicity score > 0.5 from the Helix algorithm^4^. A total of 58 variants were identified in 21 genes (Table S2). Subjects (N = 38) were excluded from the sporadic group if they had a predicted pathogenic in one of the genes.

*Analysis methods*

All statistical analyses were conducted in R (version 4.0.2). Differences between cases and controls were calculated using logistic regression including sex, age at recruitment, fasting time, years between plasma sample collection and protein analysis, and ethnicity as covariates. Survival analysis was performed using Cox proportional hazards regression with the same covariates. Time-dependent ROC calculations were performed using the timeROC package^5^. Subjects with missing data were excluded.

**Supplemental Tables**

**Table S1. Filtered subject counts**

| **Subject group** | **NfL measure** | **Prior MND diagnosis** | **Prior MND diagnosis Hospital Record** | **Prior MND diagnosis General Practitioner Record** | **Prior MND diagnosis Self Report** | **Diagnosis of G12.2 in Hospital Record** | **Evidence of exome variant in ALS gene** | **Randomly selected from UKB** | **n** |
| --- | --- | --- | --- | --- | --- | --- | --- | --- | --- |
| Filtered no NfL measure | FALSE | FALSE | FALSE | FALSE | FALSE | FALSE | FALSE | NA | 453818 |
| Filtered no NfL measure | FALSE | FALSE | FALSE | FALSE | FALSE | FALSE | TRUE | NA | 22 |
| Filtered no NfL measure | FALSE | FALSE | FALSE | FALSE | FALSE | TRUE | FALSE | NA | 24 |
| Filtered no NfL measure | FALSE | FALSE | FALSE | FALSE | FALSE | TRUE | TRUE | NA | 4 |
| Filtered no NfL measure | FALSE | TRUE | FALSE | FALSE | TRUE | FALSE | FALSE | NA | 16 |
| Filtered no NfL measure | FALSE | TRUE | FALSE | FALSE | TRUE | FALSE | TRUE | NA | 2 |
| Filtered no NfL measure | FALSE | TRUE | TRUE | FALSE | TRUE | TRUE | FALSE | NA | 1 |
| Filtered not randomly selected or case | TRUE | FALSE | FALSE | FALSE | FALSE | FALSE | FALSE | FALSE | 5560 |
| Filtered not randomly selected or case | TRUE | FALSE | FALSE | FALSE | FALSE | FALSE | TRUE | FALSE | 5 |
| Filtered prior MND | TRUE | TRUE | FALSE | FALSE | TRUE | FALSE | FALSE | FALSE | 20 |
| Filtered prior MND | TRUE | TRUE | FALSE | FALSE | TRUE | TRUE | FALSE | FALSE | 9 |
| Filtered prior MND | TRUE | TRUE | FALSE | FALSE | TRUE | TRUE | TRUE | FALSE | 2 |
| Filtered prior MND | TRUE | TRUE | TRUE | FALSE | FALSE | TRUE | FALSE | FALSE | 14 |
| Filtered prior MND | TRUE | TRUE | TRUE | FALSE | FALSE | TRUE | TRUE | FALSE | 1 |
| Filtered prior MND | TRUE | TRUE | TRUE | FALSE | TRUE | TRUE | FALSE | FALSE | 7 |
| control | TRUE | FALSE | FALSE | FALSE | FALSE | FALSE | FALSE | TRUE | 42752 |
| potential non-sporadic MND case | TRUE | FALSE | FALSE | FALSE | FALSE | TRUE | TRUE | FALSE | 32 |
| potential non-sporadic MND case | TRUE | FALSE | FALSE | FALSE | FALSE | TRUE | TRUE | TRUE | 6 |
| sporadic MND case | TRUE | FALSE | FALSE | FALSE | FALSE | TRUE | FALSE | FALSE | 184 |
| sporadic MND case | TRUE | FALSE | FALSE | FALSE | FALSE | TRUE | FALSE | TRUE | 19 |

**Table S2. List of variants considered likely deleterious in ALS genes**

| **chr_pos_ref_alt** | **Chr_b38** | **Pos_b38** | **REF** | **ALT** | **Gene_Symbol** | **Gene_ID** | **Transcript_ID** | **vep_most_severe_consequence** | **Amino_Acids** | **LOF** | **helix** |
| --- | --- | --- | --- | --- | --- | --- | --- | --- | --- | --- | --- |
| **2:201704141:A:C** | **chr2** | **201704141** | **A** | **C** | **ALS2** | **ENSG00000003393** | **ENST00000264276** | **missense_variant** | **I/R** | **null** | **TRUE** |
| **9:27548423:C:A** | **chr9** | **27548423** | **C** | **A** | **C9orf72** | **ENSG00000147894** | **ENST00000380003** | **splice_acceptor_variant** | **null** | **HC** | **FALSE** |
| **9:27548423:C:A** | **chr9** | **27548423** | **C** | **A** | **C9orf72** | **ENSG00000147894** | **ENST00000619707** | **splice_acceptor_variant** | **null** | **HC** | **FALSE** |
| **9:27548423:C:A** | **chr9** | **27548423** | **C** | **A** | **C9orf72** | **ENSG00000147894** | **ENST00000644136** | **splice_acceptor_variant** | **null** | **HC** | **FALSE** |
| **16:2437263:G:A** | **chr16** | **2437263** | **G** | **A** | **CCNF** | **ENSG00000162063** | **ENST00000397066** | **missense_variant** | **G/R** | **null** | **TRUE** |
| **12:108885136:A:C** | **chr12** | **108885136** | **A** | **C** | **DAO** | **ENSG00000110887** | **ENST00000228476** | **missense_variant** | **T/P** | **null** | **TRUE** |
| **12:108885136:A:C** | **chr12** | **108885136** | **A** | **C** | **DAO** | **ENSG00000110887** | **ENST00000547166** | **missense_variant** | **T/P** | **null** | **TRUE** |
| **12:108885136:A:C** | **chr12** | **108885136** | **A** | **C** | **DAO** | **ENSG00000110887** | **ENST00000551281** | **missense_variant** | **T/P** | **null** | **TRUE** |
| **12:108885139:A:C** | **chr12** | **108885139** | **A** | **C** | **DAO** | **ENSG00000110887** | **ENST00000228476** | **missense_variant** | **T/P** | **null** | **TRUE** |
| **12:108885139:A:C** | **chr12** | **108885139** | **A** | **C** | **DAO** | **ENSG00000110887** | **ENST00000547166** | **missense_variant** | **T/P** | **null** | **TRUE** |
| **12:108885139:A:C** | **chr12** | **108885139** | **A** | **C** | **DAO** | **ENSG00000110887** | **ENST00000551281** | **missense_variant** | **T/P** | **null** | **TRUE** |
| **2:74372936:G:A** | **chr2** | **74372936** | **G** | **A** | **DCTN1** | **ENSG00000204843** | **ENST00000361874** | **stop_gained** | **R/*** | **HC** | **FALSE** |
| **2:74372936:G:A** | **chr2** | **74372936** | **G** | **A** | **DCTN1** | **ENSG00000204843** | **ENST00000394003** | **stop_gained** | **R/*** | **HC** | **FALSE** |
| **2:74372936:G:A** | **chr2** | **74372936** | **G** | **A** | **DCTN1** | **ENSG00000204843** | **ENST00000409438** | **stop_gained** | **R/*** | **HC** | **FALSE** |
| **2:74372936:G:A** | **chr2** | **74372936** | **G** | **A** | **DCTN1** | **ENSG00000204843** | **ENST00000409868** | **stop_gained** | **R/*** | **HC** | **FALSE** |
| **2:74372936:G:A** | **chr2** | **74372936** | **G** | **A** | **DCTN1** | **ENSG00000204843** | **ENST00000458655** | **stop_gained** | **R/*** | **HC** | **FALSE** |
| **2:74372936:G:A** | **chr2** | **74372936** | **G** | **A** | **DCTN1** | **ENSG00000204843** | **ENST00000628224** | **stop_gained** | **R/*** | **HC** | **FALSE** |
| **2:74372936:G:A** | **chr2** | **74372936** | **G** | **A** | **DCTN1** | **ENSG00000204843** | **ENST00000633691** | **stop_gained** | **R/*** | **HC** | **FALSE** |
| **8:28107972:A:G** | **chr8** | **28107972** | **A** | **G** | **ELP3** | **ENSG00000134014** | **ENST00000256398** | **missense_variant** | **Y/C** | **null** | **TRUE** |
| **8:28107972:A:G** | **chr8** | **28107972** | **A** | **G** | **ELP3** | **ENSG00000134014** | **ENST00000380353** | **missense_variant** | **Y/C** | **null** | **TRUE** |
| **8:28107972:A:G** | **chr8** | **28107972** | **A** | **G** | **ELP3** | **ENSG00000134014** | **ENST00000520288** | **missense_variant** | **Y/C** | **null** | **TRUE** |
| **8:28107972:A:G** | **chr8** | **28107972** | **A** | **G** | **ELP3** | **ENSG00000134014** | **ENST00000521015** | **missense_variant** | **Y/C** | **null** | **TRUE** |
| **8:28107972:A:G** | **chr8** | **28107972** | **A** | **G** | **ELP3** | **ENSG00000134014** | **ENST00000521570** | **missense_variant** | **Y/C** | **null** | **TRUE** |
| **8:28107972:A:G** | **chr8** | **28107972** | **A** | **G** | **ELP3** | **ENSG00000134014** | **ENST00000524103** | **missense_variant** | **Y/C** | **null** | **TRUE** |
| **6:109715133:T:C** | **chr6** | **109715133** | **T** | **C** | **FIG4** | **ENSG00000112367** | **ENST00000230124** | **missense_variant** | **I/T** | **null** | **TRUE** |
| **6:109715133:T:C** | **chr6** | **109715133** | **T** | **C** | **FIG4** | **ENSG00000112367** | **ENST00000454215** | **missense_variant** | **I/T** | **null** | **TRUE** |
| **6:109716560:G:A** | **chr6** | **109716560** | **G** | **A** | **FIG4** | **ENSG00000112367** | **ENST00000230124** | **missense_variant** | **G/D** | **null** | **TRUE** |
| **6:109716560:G:A** | **chr6** | **109716560** | **G** | **A** | **FIG4** | **ENSG00000112367** | **ENST00000368941** | **missense_variant** | **G/D** | **null** | **TRUE** |
| **6:109716560:G:A** | **chr6** | **109716560** | **G** | **A** | **FIG4** | **ENSG00000112367** | **ENST00000454215** | **missense_variant** | **G/D** | **null** | **TRUE** |
| **9:36217448:C:A** | **chr9** | **36217448** | **C** | **A** | **GNE** | **ENSG00000159921** | **ENST00000377902** | **missense_variant** | **V/L** | **null** | **TRUE** |
| **9:36217448:C:A** | **chr9** | **36217448** | **C** | **A** | **GNE** | **ENSG00000159921** | **ENST00000396594** | **missense_variant** | **V/L** | **null** | **TRUE** |
| **9:36217448:C:A** | **chr9** | **36217448** | **C** | **A** | **GNE** | **ENSG00000159921** | **ENST00000447283** | **missense_variant** | **V/L** | **null** | **TRUE** |
| **9:36217448:C:A** | **chr9** | **36217448** | **C** | **A** | **GNE** | **ENSG00000159921** | **ENST00000539208** | **missense_variant** | **V/L** | **null** | **TRUE** |
| **9:36217448:C:A** | **chr9** | **36217448** | **C** | **A** | **GNE** | **ENSG00000159921** | **ENST00000539815** | **missense_variant** | **V/L** | **null** | **TRUE** |
| **9:36217448:C:A** | **chr9** | **36217448** | **C** | **A** | **GNE** | **ENSG00000159921** | **ENST00000543356** | **missense_variant** | **V/L** | **null** | **TRUE** |
| **9:36217448:C:A** | **chr9** | **36217448** | **C** | **A** | **GNE** | **ENSG00000159921** | **ENST00000642385** | **missense_variant** | **V/L** | **null** | **TRUE** |
| **9:36227397:C:A** | **chr9** | **36227397** | **C** | **A** | **GNE** | **ENSG00000159921** | **ENST00000377902** | **missense_variant** | **D/Y** | **null** | **TRUE** |
| **9:36227397:C:A** | **chr9** | **36227397** | **C** | **A** | **GNE** | **ENSG00000159921** | **ENST00000396594** | **missense_variant** | **D/Y** | **null** | **TRUE** |
| **9:36227397:C:A** | **chr9** | **36227397** | **C** | **A** | **GNE** | **ENSG00000159921** | **ENST00000447283** | **missense_variant** | **D/Y** | **null** | **TRUE** |
| **9:36227397:C:A** | **chr9** | **36227397** | **C** | **A** | **GNE** | **ENSG00000159921** | **ENST00000539208** | **missense_variant** | **D/Y** | **null** | **TRUE** |
| **9:36227397:C:A** | **chr9** | **36227397** | **C** | **A** | **GNE** | **ENSG00000159921** | **ENST00000539815** | **missense_variant** | **D/Y** | **null** | **TRUE** |
| **9:36227397:C:A** | **chr9** | **36227397** | **C** | **A** | **GNE** | **ENSG00000159921** | **ENST00000543356** | **missense_variant** | **D/Y** | **null** | **TRUE** |
| **9:36227397:C:A** | **chr9** | **36227397** | **C** | **A** | **GNE** | **ENSG00000159921** | **ENST00000642385** | **missense_variant** | **D/Y** | **null** | **TRUE** |
| **9:36246049:T:A** | **chr9** | **36246049** | **T** | **A** | **GNE** | **ENSG00000159921** | **ENST00000377902** | **missense_variant** | **I/F** | **null** | **TRUE** |
| **9:36246049:T:A** | **chr9** | **36246049** | **T** | **A** | **GNE** | **ENSG00000159921** | **ENST00000396594** | **missense_variant** | **I/F** | **null** | **TRUE** |
| **9:36246049:T:A** | **chr9** | **36246049** | **T** | **A** | **GNE** | **ENSG00000159921** | **ENST00000447283** | **missense_variant** | **I/F** | **null** | **TRUE** |
| **9:36246049:T:A** | **chr9** | **36246049** | **T** | **A** | **GNE** | **ENSG00000159921** | **ENST00000539208** | **missense_variant** | **I/F** | **null** | **TRUE** |
| **9:36246049:T:A** | **chr9** | **36246049** | **T** | **A** | **GNE** | **ENSG00000159921** | **ENST00000539815** | **missense_variant** | **I/F** | **null** | **TRUE** |
| **9:36246049:T:A** | **chr9** | **36246049** | **T** | **A** | **GNE** | **ENSG00000159921** | **ENST00000543356** | **missense_variant** | **I/F** | **null** | **TRUE** |
| **9:36246049:T:A** | **chr9** | **36246049** | **T** | **A** | **GNE** | **ENSG00000159921** | **ENST00000642385** | **missense_variant** | **I/F** | **null** | **TRUE** |
| **9:36276897:A:T** | **chr9** | **36276897** | **A** | **T** | **GNE** | **ENSG00000159921** | **ENST00000543356** | **missense_variant** | **L/H** | **null** | **TRUE** |
| **12:57569671:C:T** | **chr12** | **57569671** | **C** | **T** | **KIF5A** | **ENSG00000155980** | **ENST00000286452** | **missense_variant** | **R/W** | **null** | **TRUE** |
| **12:57569671:C:T** | **chr12** | **57569671** | **C** | **T** | **KIF5A** | **ENSG00000155980** | **ENST00000455537** | **missense_variant** | **R/W** | **null** | **TRUE** |
| **12:57581151:C:T** | **chr12** | **57581151** | **C** | **T** | **KIF5A** | **ENSG00000155980** | **ENST00000286452** | **missense_variant** | **R/W** | **null** | **TRUE** |
| **12:57581151:C:T** | **chr12** | **57581151** | **C** | **T** | **KIF5A** | **ENSG00000155980** | **ENST00000455537** | **missense_variant** | **R/W** | **null** | **TRUE** |
| **5:126804834:G:C** | **chr5** | **126804834** | **G** | **C** | **LMNB1** | **ENSG00000113368** | **ENST00000261366** | **missense_variant** | **A/P** | **null** | **TRUE** |
| **5:126804834:G:C** | **chr5** | **126804834** | **G** | **C** | **LMNB1** | **ENSG00000113368** | **ENST00000395354** | **missense_variant** | **A/P** | **null** | **TRUE** |
| **5:126804834:G:C** | **chr5** | **126804834** | **G** | **C** | **LMNB1** | **ENSG00000113368** | **ENST00000492190** | **missense_variant** | **A/P** | **null** | **TRUE** |
| **5:139308062:G:C** | **chr5** | **139308062** | **G** | **C** | **MATR3** | **ENSG00000280987** | **ENST00000361059** | **missense_variant** | **R/T** | **null** | **TRUE** |
| **5:139308062:G:C** | **chr5** | **139308062** | **G** | **C** | **MATR3** | **ENSG00000280987** | **ENST00000394800** | **missense_variant** | **R/T** | **null** | **TRUE** |
| **5:139308062:G:C** | **chr5** | **139308062** | **G** | **C** | **MATR3** | **ENSG00000015479** | **ENST00000394805** | **missense_variant** | **R/T** | **null** | **TRUE** |
| **5:139308062:G:C** | **chr5** | **139308062** | **G** | **C** | **MATR3** | **ENSG00000280987** | **ENST00000502929** | **missense_variant** | **R/T** | **null** | **TRUE** |
| **5:139308062:G:C** | **chr5** | **139308062** | **G** | **C** | **MATR3** | **ENSG00000015479** | **ENST00000504045** | **missense_variant** | **R/T** | **null** | **TRUE** |
| **5:139308062:G:C** | **chr5** | **139308062** | **G** | **C** | **MATR3** | **ENSG00000280987** | **ENST00000509990** | **missense_variant** | **R/T** | **null** | **TRUE** |
| **5:139308062:G:C** | **chr5** | **139308062** | **G** | **C** | **MATR3** | **ENSG00000015479** | **ENST00000510056** | **missense_variant** | **R/T** | **null** | **TRUE** |
| **5:139308062:G:C** | **chr5** | **139308062** | **G** | **C** | **MATR3** | **ENSG00000015479** | **ENST00000618441** | **missense_variant** | **R/T** | **null** | **TRUE** |
| **22:29485808:A:C** | **chr22** | **29485808** | **A** | **C** | **NEFH** | **ENSG00000100285** | **ENST00000310624** | **missense_variant** | **N/T** | **null** | **TRUE** |
| **4:169580841:C:G** | **chr4** | **169580841** | **C** | **G** | **NEK1** | **ENSG00000137601** | **ENST00000439128** | **splice_donor_variant** | **null** | **HC** | **FALSE** |
| **4:169580841:C:G** | **chr4** | **169580841** | **C** | **G** | **NEK1** | **ENSG00000137601** | **ENST00000505119** | **splice_donor_variant** | **null** | **HC** | **FALSE** |
| **4:169580841:C:G** | **chr4** | **169580841** | **C** | **G** | **NEK1** | **ENSG00000137601** | **ENST00000507142** | **splice_donor_variant** | **null** | **HC** | **FALSE** |
| **4:169580841:C:G** | **chr4** | **169580841** | **C** | **G** | **NEK1** | **ENSG00000137601** | **ENST00000510533** | **splice_donor_variant** | **null** | **HC** | **FALSE** |
| **4:169580841:C:G** | **chr4** | **169580841** | **C** | **G** | **NEK1** | **ENSG00000137601** | **ENST00000511633** | **splice_donor_variant** | **null** | **HC** | **FALSE** |
| **4:169580841:C:G** | **chr4** | **169580841** | **C** | **G** | **NEK1** | **ENSG00000137601** | **ENST00000512193** | **splice_donor_variant** | **null** | **HC** | **FALSE** |
| **4:169587593:C:T** | **chr4** | **169587593** | **C** | **T** | **NEK1** | **ENSG00000137601** | **ENST00000439128** | **missense_variant** | **C/Y** | **null** | **TRUE** |
| **4:169587593:C:T** | **chr4** | **169587593** | **C** | **T** | **NEK1** | **ENSG00000137601** | **ENST00000507142** | **missense_variant** | **C/Y** | **null** | **TRUE** |
| **4:169587593:C:T** | **chr4** | **169587593** | **C** | **T** | **NEK1** | **ENSG00000137601** | **ENST00000510533** | **missense_variant** | **C/Y** | **null** | **TRUE** |
| **4:169587593:C:T** | **chr4** | **169587593** | **C** | **T** | **NEK1** | **ENSG00000137601** | **ENST00000511633** | **missense_variant** | **C/Y** | **null** | **TRUE** |
| **4:169587593:C:T** | **chr4** | **169587593** | **C** | **T** | **NEK1** | **ENSG00000137601** | **ENST00000512193** | **missense_variant** | **C/Y** | **null** | **TRUE** |
| **4:169589477:C:T** | **chr4** | **169589477** | **C** | **T** | **NEK1** | **ENSG00000137601** | **ENST00000439128** | **missense_variant** | **G/E** | **null** | **TRUE** |
| **4:169589477:C:T** | **chr4** | **169589477** | **C** | **T** | **NEK1** | **ENSG00000137601** | **ENST00000507142** | **missense_variant** | **G/E** | **null** | **TRUE** |
| **4:169589477:C:T** | **chr4** | **169589477** | **C** | **T** | **NEK1** | **ENSG00000137601** | **ENST00000510533** | **missense_variant** | **G/E** | **null** | **TRUE** |
| **4:169589477:C:T** | **chr4** | **169589477** | **C** | **T** | **NEK1** | **ENSG00000137601** | **ENST00000511633** | **missense_variant** | **G/E** | **null** | **TRUE** |
| **4:169589477:C:T** | **chr4** | **169589477** | **C** | **T** | **NEK1** | **ENSG00000137601** | **ENST00000512193** | **missense_variant** | **G/E** | **null** | **TRUE** |
| **4:169602600:C:T** | **chr4** | **169602600** | **C** | **T** | **NEK1** | **ENSG00000137601** | **ENST00000439128** | **missense_variant** | **G/R** | **null** | **TRUE** |
| **4:169602600:C:T** | **chr4** | **169602600** | **C** | **T** | **NEK1** | **ENSG00000137601** | **ENST00000507142** | **missense_variant** | **G/R** | **null** | **TRUE** |
| **4:169602600:C:T** | **chr4** | **169602600** | **C** | **T** | **NEK1** | **ENSG00000137601** | **ENST00000510533** | **missense_variant** | **G/R** | **null** | **TRUE** |
| **4:169602600:C:T** | **chr4** | **169602600** | **C** | **T** | **NEK1** | **ENSG00000137601** | **ENST00000511633** | **missense_variant** | **G/R** | **null** | **TRUE** |
| **4:169602600:C:T** | **chr4** | **169602600** | **C** | **T** | **NEK1** | **ENSG00000137601** | **ENST00000512193** | **missense_variant** | **G/R** | **null** | **TRUE** |
| **12:49297274:G:A** | **chr12** | **49297274** | **G** | **A** | **PRPH** | **ENSG00000135406** | **ENST00000257860** | **splice_donor_variant** | **null** | **HC** | **FALSE** |
| **12:49297274:G:A** | **chr12** | **49297274** | **G** | **A** | **PRPH** | **ENSG00000135406** | **ENST00000532332** | **splice_donor_variant** | **null** | **HC** | **FALSE** |
| **21:31667359:T:C** | **chr21** | **31667359** | **T** | **C** | **SOD1** | **ENSG00000142168** | **ENST00000270142** | **missense_variant** | **I/T** | **null** | **TRUE** |
| **21:31667359:T:C** | **chr21** | **31667359** | **T** | **C** | **SOD1** | **ENSG00000142168** | **ENST00000389995** | **missense_variant** | **I/T** | **null** | **TRUE** |
| **15:44583835:GA:G** | **chr15** | **44583835** | **GA** | **G** | **SPG11** | **ENSG00000104133** | **ENST00000261866** | **frameshift_variant** | **I/X** | **HC** | **FALSE** |
| **15:44583835:GA:G** | **chr15** | **44583835** | **GA** | **G** | **SPG11** | **ENSG00000104133** | **ENST00000427534** | **frameshift_variant** | **I/X** | **HC** | **FALSE** |
| **15:44583835:GA:G** | **chr15** | **44583835** | **GA** | **G** | **SPG11** | **ENSG00000104133** | **ENST00000535302** | **frameshift_variant** | **I/X** | **HC** | **FALSE** |
| **15:44583835:GA:G** | **chr15** | **44583835** | **GA** | **G** | **SPG11** | **ENSG00000104133** | **ENST00000558319** | **frameshift_variant** | **I/X** | **HC** | **FALSE** |
| **15:44583835:GA:G** | **chr15** | **44583835** | **GA** | **G** | **SPG11** | **ENSG00000104133** | **ENST00000559511** | **frameshift_variant** | **I/X** | **HC** | **FALSE** |
| **5:179836445:C:T** | **chr5** | **179836445** | **C** | **T** | **SQSTM1** | **ENSG00000161011** | **ENST00000360718** | **missense_variant** | **P/L** | **null** | **TRUE** |
| **5:179836445:C:T** | **chr5** | **179836445** | **C** | **T** | **SQSTM1** | **ENSG00000161011** | **ENST00000389805** | **missense_variant** | **P/L** | **null** | **TRUE** |
| **5:179836453:A:AT** | **chr5** | **179836453** | **A** | **AT** | **SQSTM1** | **ENSG00000161011** | **ENST00000360718** | **frameshift_variant** | **I/IX** | **HC** | **FALSE** |
| **5:179836453:A:AT** | **chr5** | **179836453** | **A** | **AT** | **SQSTM1** | **ENSG00000161011** | **ENST00000389805** | **frameshift_variant** | **I/IX** | **HC** | **FALSE** |
| **12:64497979:TA:T** | **chr12** | **64497979** | **TA** | **T** | **TBK1** | **ENSG00000183735** | **ENST00000331710** | **frameshift_variant** | **L/X** | **HC** | **FALSE** |
| **12:64497979:TA:T** | **chr12** | **64497979** | **TA** | **T** | **TBK1** | **ENSG00000183735** | **ENST00000650762** | **frameshift_variant** | **L/X** | **HC** | **FALSE** |
| **12:64497979:TA:T** | **chr12** | **64497979** | **TA** | **T** | **TBK1** | **ENSG00000183735** | **ENST00000650790** | **frameshift_variant** | **L/X** | **HC** | **FALSE** |
| **12:64497979:TA:T** | **chr12** | **64497979** | **TA** | **T** | **TBK1** | **ENSG00000183735** | **ENST00000651014** | **frameshift_variant** | **L/X** | **HC** | **FALSE** |
| **2:219250546:C:T** | **chr2** | **219250546** | **C** | **T** | **TUBA4A** | **ENSG00000127824** | **ENST00000248437** | **missense_variant** | **A/T** | **null** | **TRUE** |
| **2:219250546:C:T** | **chr2** | **219250546** | **C** | **T** | **TUBA4A** | **ENSG00000127824** | **ENST00000392088** | **missense_variant** | **A/T** | **null** | **TRUE** |
| **19:17632876:C:T** | **chr19** | **17632876** | **C** | **T** | **UNC13A** | **ENSG00000130477** | **ENST00000519716** | **missense_variant** | **D/N** | **null** | **TRUE** |
| **19:17632876:C:T** | **chr19** | **17632876** | **C** | **T** | **UNC13A** | **ENSG00000130477** | **ENST00000550896** | **missense_variant** | **D/N** | **null** | **TRUE** |
| **19:17632876:C:T** | **chr19** | **17632876** | **C** | **T** | **UNC13A** | **ENSG00000130477** | **ENST00000551649** | **missense_variant** | **D/N** | **null** | **TRUE** |
| **19:17632876:C:T** | **chr19** | **17632876** | **C** | **T** | **UNC13A** | **ENSG00000130477** | **ENST00000552293** | **missense_variant** | **D/N** | **null** | **TRUE** |
| **9:35067916:G:A** | **chr9** | **35067916** | **G** | **A** | **VCP** | **ENSG00000165280** | **ENST00000358901** | **missense_variant** | **R/C** | **null** | **TRUE** |
| **9:35067916:G:A** | **chr9** | **35067916** | **G** | **A** | **VCP** | **ENSG00000165280** | **ENST00000417448** | **missense_variant** | **R/C** | **null** | **TRUE** |
| **9:35067916:G:A** | **chr9** | **35067916** | **G** | **A** | **VCP** | **ENSG00000165280** | **ENST00000448530** | **missense_variant** | **R/C** | **null** | **TRUE** |
| **9:35068264:A:C** | **chr9** | **35068264** | **A** | **C** | **VCP** | **ENSG00000165280** | **ENST00000358901** | **missense_variant** | **V/G** | **null** | **TRUE** |

**Table S3: Survival association and AUC results for other neurodegenerative diseases**

| **Diagnosis** | **estimate** | **std.error** | **p.value** | **time** | **Cases** | **survivor at t** | **Censored at t** | **AUC** |
| --- | --- | --- | --- | --- | --- | --- | --- | --- |
| G20 Parkinson's disease | 0.402 | 0.064 | 2.98E-10 | t=1 | 28 | 35352 | 911 | 0.77 |
| G20 Parkinson's disease | 0.402 | 0.064 | 2.98E-10 | t=2 | 85 | 34241 | 1965 | 0.67 |
| G20 Parkinson's disease | 0.402 | 0.064 | 2.98E-10 | t=3 | 157 | 32963 | 3171 | 0.66 |
| G20 Parkinson's disease | 0.402 | 0.064 | 2.98E-10 | t=5 | 392 | 29792 | 6107 | 0.66 |
| G20 Parkinson's disease | 0.402 | 0.064 | 2.98E-10 | t=10 | 650 | 13961 | 21680 | 0.65 |
| G30 Alzheimer's disease | 0.752 | 0.068 | 4.69E-28 | t=1 | 3 | 35411 | 913 | 0.75 |
| G30 Alzheimer's disease | 0.752 | 0.068 | 4.69E-28 | t=2 | 13 | 34347 | 1967 | 0.77 |
| G30 Alzheimer's disease | 0.752 | 0.068 | 4.69E-28 | t=3 | 29 | 33119 | 3179 | 0.76 |
| G30 Alzheimer's disease | 0.752 | 0.068 | 4.69E-28 | t=5 | 85 | 30093 | 6149 | 0.81 |
| G30 Alzheimer's disease | 0.752 | 0.068 | 4.69E-28 | t=10 | 349 | 14114 | 21864 | 0.75 |
| G31.0 Circumscribed brain atrophy | 1.100 | 0.110 | 1.97E-23 | t=1 | 1 | 35397 | 914 | 0.85 |
| G31.0 Circumscribed brain atrophy | 1.100 | 0.110 | 1.97E-23 | t=2 | 2 | 34340 | 1970 | 0.87 |
| G31.0 Circumscribed brain atrophy | 1.100 | 0.110 | 1.97E-23 | t=3 | 5 | 33122 | 3185 | 0.91 |
| G31.0 Circumscribed brain atrophy | 1.100 | 0.110 | 1.97E-23 | t=5 | 23 | 30125 | 6164 | 0.89 |
| G31.0 Circumscribed brain atrophy | 1.100 | 0.110 | 1.97E-23 | t=10 | 84 | 14221 | 22007 | 0.76 |

**Supplemental Figures**

**Figure S1.** NfL levels vs. time from diagnosis for MND cases stratified by carrier status for ALS familial risk genes. Scatterplots showing NfL expression vs. time from MND diagnosis including subjects with likely deleterious variants in ALS genes. A) Point colors indicate the gene that was reported with a likely deleterious variant for that participant. B) The black and red solid lines indicates smoothed lines calculated using the loess method for those with and without likely deleterious variants.

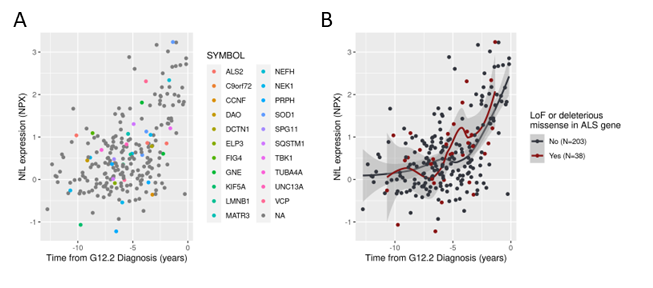

**Figure S2**. NfL levels vs. time from diagnosis for selected neurodegenerative diseases. Scatterplots showing NfL expression vs. time from diagnosis for participants with diagnoses of A) Parkinson’s disease, B) Alzheimer’s disease, or C) circumscribed brain injury, which includes frontotemporal dementia.

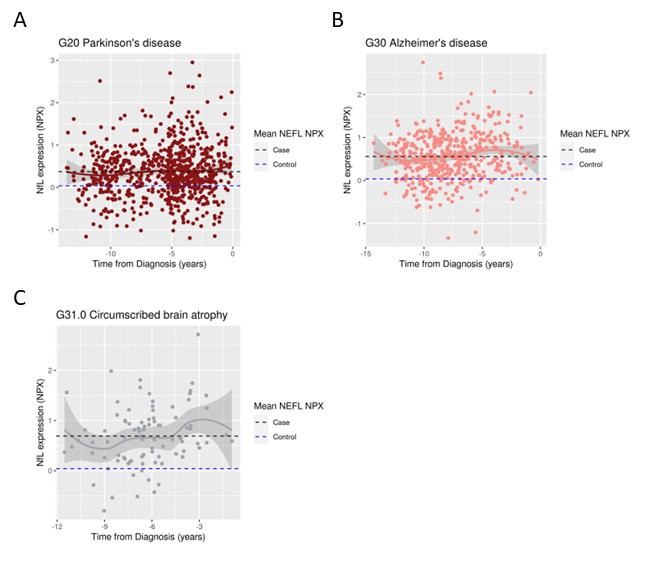

**Figure S3.** Hand grip strength vs. time from diagnosis for sporadic MND cases. Scatterplot showing values of hand grip strength for the left (left) and right (right) hands relative to the time from motor neuron disease diagnosis for sporadic MND participants. Points are colored according to sex and sex-specific smoothed lines are drawn using the loess method.
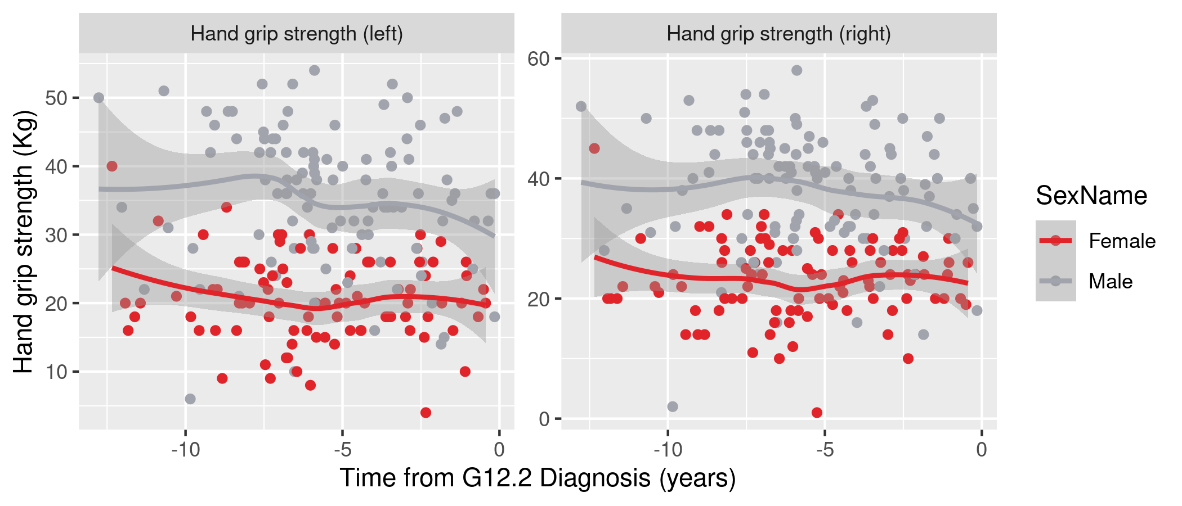

1.	Ollier W, Sprosen T, Peakman T. UK Biobank: from concept to reality. Pharmacogenomics 2005;6:639-646.

2.	Sun BB, Chiou J, Traylor M, et al. Genetic regulation of the human plasma proteome in 54,306 UK Biobank participants. bioRxiv 2022.

3.	Mejzini R, Flynn LL, Pitout IL, Fletcher S, Wilton SD, Akkari PA. ALS Genetics, Mechanisms, and Therapeutics: Where Are We Now? Front Neurosci 2019;13:1310.

4.	Vroling B, Heijl S. White paper: the helix pathogenicity prediction platform. arXiv preprint arXiv:210401033 2021.

5.	Blanche P, Dartigues JF, Jacqmin-Gadda H. Estimating and comparing time-dependent areas under receiver operating characteristic curves for censored event times with competing risks. Stat Med 2013;32:5381-5397.
